# Predicting Air Conditioner Ownership in Low- and Middle-Income Countries using Geospatial Machine Learning Methods - Mapping Cooling Inequality

**DOI:** 10.64898/2026.10.12.26365281

**Authors:** Amber Vayani, Simon E. H. Smart, Tim CD Lucas

**Affiliations:** Biostatistics Research Group, University of Leicester.

## Abstract

**Background:** Rising global temperatures are intensifying exposure to extreme heat, yet access to air conditioning (AC) remains unequal which leaves vulnerable populations without protection against the heat. Household survey data on AC ownership is scarce in low- and middle-income countries limiting the ability to identify areas of cooling gaps.

**Methods:** In this study, we develop a geospatial machine learning framework that integrates household data from the Demographic and Health Surveys (DHS) program with high resolution climatic, socioeconomic and demographic covariates to generate a 5km resolution prediction of AC ownership across low- and middle-income countries in Africa and Asia for the year 2023.

**Results:** The resulting map reveals strong spatial inequalities with high predicted prevalence in wealthier and urban regions, and near zero prevalence across much of Central and West Africa and rural areas. These patterns reflect the adaptation cooling gap where those most exposed to extreme heat are least able to adapt through access to cooling.

**Conclusion:** By producing the first continental-scale, high resolution map of AC ownership, this study lays the foundation for designing and targeting equitable cooling strategies, support heat-health planning particularly in communities most affected and informing sustainable energy policy.

## Introduction

Rising global temperatures are intensifying the extent of extreme heat. Global organisations such as Intergovernmental Panel on Climate Change (IPCC) and the World Health Organisation (WHO) recognise this as one of the most severe climate related hazards. Global mean surface temperatures have already risen by approximately 1.1 °C since pre-industrial times, primarily due to anthropogenic greenhouse gas emissions (IPCC 2021). Heatwaves are becoming more frequent, prolonged and severe, especially in tropical and subtropical regions (Meher et al. 2025; WHO 2024). Projections under current emission scenarios suggest that without rapid climate action, both the scale and intensity of heat exposure will continue to escalate over the coming decades (Lindsey and Dahlman 2025).

Extreme heat has direct physiological effects like dehydration, heat exhaustion and heat stroke as well as public health implications (Faurie et al. 2022). For example, in the 2003 European heatwave, more than 50% of heat-related deaths occurred indoors, disproportionately affecting the elderly and those with pre-existing conditions (Ballester et al. 2023; Ahn and Uejio 2022).

Current WHO estimates suggest that heatwaves are already responsible for nearly half a million deaths globally annually, and is expected to rise significantly as climate continues to warm (WHO, 2024). Children, the elderly, people with pre-existing comorbidities and low income households are especially vulnerable due to physiological sensitivity and limited adaptive capacity (Campbell et al. 2018; Howard and Krishna 2022).

Air conditioning (AC) is one of the most effective technological adaptations to protect human health from the dangers of excessive heat. AC reduces heat-related mortality and enhances comfort, sleep quality and productivity (Zhang et al. 2024). The International Energy Agency (IEA) estimates that by 2050, two thirds of the world’s households could own an AC unit, up from a current value of about 1.6 billion units (IEA 2018; Zhang et al. 2024). Demand is growing fastest in low- and middle-income countries (LMICs), many of which are entering the steep phase of the income-driven S-curve of AC adoption (Pavanello et al. 2021; Davis et al. 2021).

However, access to AC remains unequal. Despite the rising need, populations in the hottest regions still lack adequate access to AC due to affordability barriers, poor housing infrastructure and unreliable electricity (Davis et al. 2021; Pavanello et al. 2021).This disparity is referred to as the cooling gap and reflects broader socio-economic patterns which expose marginalised groups to health risks of extreme heat (Gronlund 2014; IEA 2018). Rural communities face limited access to AC due to poverty and weak infrastructure, while wealthier urban households are rapidly increasing AC use which widens this inequality. Davis (et al. 2021) describes this as the “adaptation cooling gap”, highlighting the mismatch between populations most exposed to heat stress and those able to adapt through access to cooling. As a result, those at greatest risk of heat-related illness are often least able to access AC, thus reinforcing the cycle of health inequality.

Cooling technologies such as AC are energy intensive and remain largely dependent on fossil fuels, thus contributing to peak electricity demand, greenhouse gas emissions and localised urban heat island effects (Mohajerani et al. 2017; Zhai et al. 2024). This creates a paradox whereby AC offers protection from heat but simultaneously intensifies the drivers of global warming and directly increases local temperatures. Therefore, without careful planning, rapid AC expansion could undermine long-term climate goals and widen inequalities through rising energy poverty (Khosla 2020).

Therefore, understanding who has access to AC is important for public health researchers, urban planners and policymakers to design equitable and sustainable cooling strategies. Accurate data on AC ownership is essential for identifying vulnerability hotspots, allocating resources such as cooling centres or subsidies and targeting climate adaptation efforts where they are needed the most. However, high resolution data on AC ownership remains scarce in many LMICs. Household surveys such as the Demographic and Health Surveys (DHS) provide valuable information on household infrastructure and utility, population demographics, health and geographic information to link survey metrics with local infrastructure and environmental conditions. However, this is often geographically incomplete as geolocation data are unavailable or due to privacy restrictions that limit Global Positioning System (GPS) disclosure. This introduces spatial gaps that weaken analysis and prediction.

This study addresses this knowledge gap by developing a machine learning framework to predict household AC ownership across Africa and Asia. By integrating geolocated DHS data with remotely sensed environmental and socioeconomic variables, we produce high-resolution, cross-validated estimates of AC ownership in regions where traditional data sources are unavailable. This map of AC ownership can inform the design of equitable heat adaptation policies and contribute towards future epidemiological studies on the protective and adverse effects of AC on human health.

## Methods

We identified national household surveys from the DHS programme that contained both household AC ownership and geographic coordinates from the year 2000 onwards using standard recode files (ICF 2003-2023). Datasets from all 24 eligible countries in Africa and Asia were merged, with the primary outcome being the proportion of households in a cluster with AC (Figure 4).

To complement survey data, we obtained high-resolution (5 km at the equator) global raster datasets representing environmental, climatic and socioeconomic conditions. As our aim was to make predictions across entire continents, only covariates available as global rasters were used. Therefore, no household or individual level data from the DHS surveys were used despite their predictive value. All datasets were projected to the WGS84 coordinate reference system (EPSG:4326) and indexed by year to allow temporal matching with DHS survey years.

Covariates included remotely sensed land surface temperature (day and night), diurnal temperature range, mean, maximum and minimum temperatures, precipitation, rainfall, frost days, vapour pressure, wet days, potential evapotranspiration, vegetation index and cloud cover. Additionally, the socioeconomic indicators urban-rural classification, human footprint, population density, travel time to the nearest city and nightlight composite were obtained. National level gross domestic product (GDP) was also included. Longitude and latitude were included as covariates to account for spatial structure while survey year was included as a covariate to account for changes through time. The urban-rural classification was derived from the Urban-Rural Catchment Areas (URCA) raster dataset which assigns pixels to one of 30 categories based on travel times to the nearest urban centre and size of the city. We re-coded into four ordered levels: urban (within urban centres), peri-urban (< 1 hour travel time), rural near (1-2 hours) and remote (>2 hours) as per the raster description file. All rasters were aggregated to an annual temporal resolution. A summary of these variables is in Table 4.

Household cluster coordinates from DHS were used to extract covariate values from raster layers corresponding to the year of each survey. Where the exact year was not available, the nearest year was used, maximising both spatial and temporal alignment between survey observations and environmental variables. Most climatic covariates were available annually across the DHS survey period, while others had limited coverage e.g., nightlights from 2012 onwards, population density and vegetation index up to 2020, and travel times as a single layer. These were matched to surveys using the closest available year. Small amounts of missing raster-derived covariate values arising from incomplete spatial coverage or limited temporal availability were handled by a complete-case analysis during model fitting. To reduce skewness and improve model performance, log transformations were applied to highly skewed variables including GDP, human footprint, nightlights, population density, rainfall precipitation and travel times.

We used R (v 4.5.1) for all analyses and models were fitted on ALICE at the University of Leicester. Model development began with three models implemented using the caret package; Elastic net, decision trees and random forests. Models were trained using a 10-fold cross validation repeated 10 times with complete cases only.

Model performance was evaluated using the Mean Absolute Error (MAE) between predicted and observed proportion of households with AC was the primary performance metric. Given that our unit of interest is the cluster, not the household, this cluster level performance metric is more appropriate than, for example, accuracy on a prediction of household AC ownership. Similarly, due to this focus, the elastic net was fitted on cluster level proportions with a linear link function rather than to household AC with a logistic link function. Predicted values below zero were adjusted to zero.

Random forest models outperformed the other approaches and were further optimised by tuning the hyperparameters. We conducted a repeated 10-fold cross validation with 5 repeats using a manually defined tuning grid for the number of variables sampled at each split (*mtry* = 3,6,9,12,15,18) and the minimum node size (*min*. *node*. *size* = 25,50,100). The model was weighted using DHS cluster size representing the number of households per cluster as the weight. As the outcome was continuous, variance was chosen as the split rule.

We additionally implemented a leave-country-out cross validation framework to assess model generalisability. For each iteration, data from one country was withheld as the test set, and the model was trained on all remaining countries using the same weights as in the previous model. This approach tested the model’s ability to predict AC ownership in unseen contexts, reflecting real-world scenarios where some countries do not have any household data.

To generate predictions of AC prevalence the model was applied to covariates for 2023 and masked to produce a continuous surface of predicted AC ownership across low- and middle-income countries in Africa and Asia. These predictions are available at 5km resolution at the equator.

To estimate the number of people living in extreme heat without AC in low and middle countries in Africa and Asia, we combined gridded population data and annual maximum temperature. Annual maximum temperature rasters from 2014 to 2023 were used to account for the yearly variance in exposure to extreme heat, and areas where maximum temperature exceeded 35°*C* and 40°*C* were identified. These exposure rasters were combined with gridded population count from World Pop data (Bondarenko et al. 2025) and predicted air conditioning prevalence. The proportion of households without AC was calculated as 1 − *predicted AC prevalence*, and multiplied by population counts to estimate the number of people lacking access to cooling within each grid cell. These were then summed across all exposed cells and summarised across the 2014 to 2023 period to provide an indication of the range of potential exposure under varying yearly temperature conditions.

To better understand the poor leave country out cross validation performance, we conducted additional country-level analysis. For each hold-out country, we compared the observed and predicted AC prevalence per cluster and calculated correlations between them. This was testing the model’s ability to detect which locations within the country have higher AC, and if the ranks of clusters are preserved in predictions. We also calculated the observed and predicted mean AC prevalence to highlight any errors in estimating country-level averages compared to ranking locations within countries. This was testing the model’s ability to know which countries have higher AC overall.

## Results

### Exploratory analysis

To understand how each variable relates to AC prevalence, we plotted smoothed scatter plots of each predictor against observed AC prevalence (Figure 2). These showed positive associations between AC ownership and GDP, nightlights and global human footprint capturing economic activity and infrastructure. AC prevalence also increased with temperature indicators such as day and night LST, mean temperature, minimum and maximum temperature, consistent with greater need for cooling in hotter areas.

Conversely, travel times to cities were negatively associated with AC ownership indicating lower access in remote areas. Rainfall, precipitation and wet days also showed slight negative associations with AC. Variables such as vapour pressure, vegetation index and cloud cover displayed minimal associations.

Pearson pairwise correlations among all predictors and between each predictor and the outcome were examined to assess multicollinearity and further understand variable associations (Figure 1). Correlations between predictors were generally moderate, with the strongest being between temperature (mean, max, min) and between GDP and nightlights. The variable most strongly correlated with AC ownership was maximum temperature (*r* = 0.39), nightlights (*r* = 0.36), followed by human footprint (*r* = 0.35), population density (*r* = 0.28) and diurnal temperature range (*r* = 0.25). These results suggest that economic activity, urbanisation and climate are all important predictors of AC prevalence.

**Figure 1:**
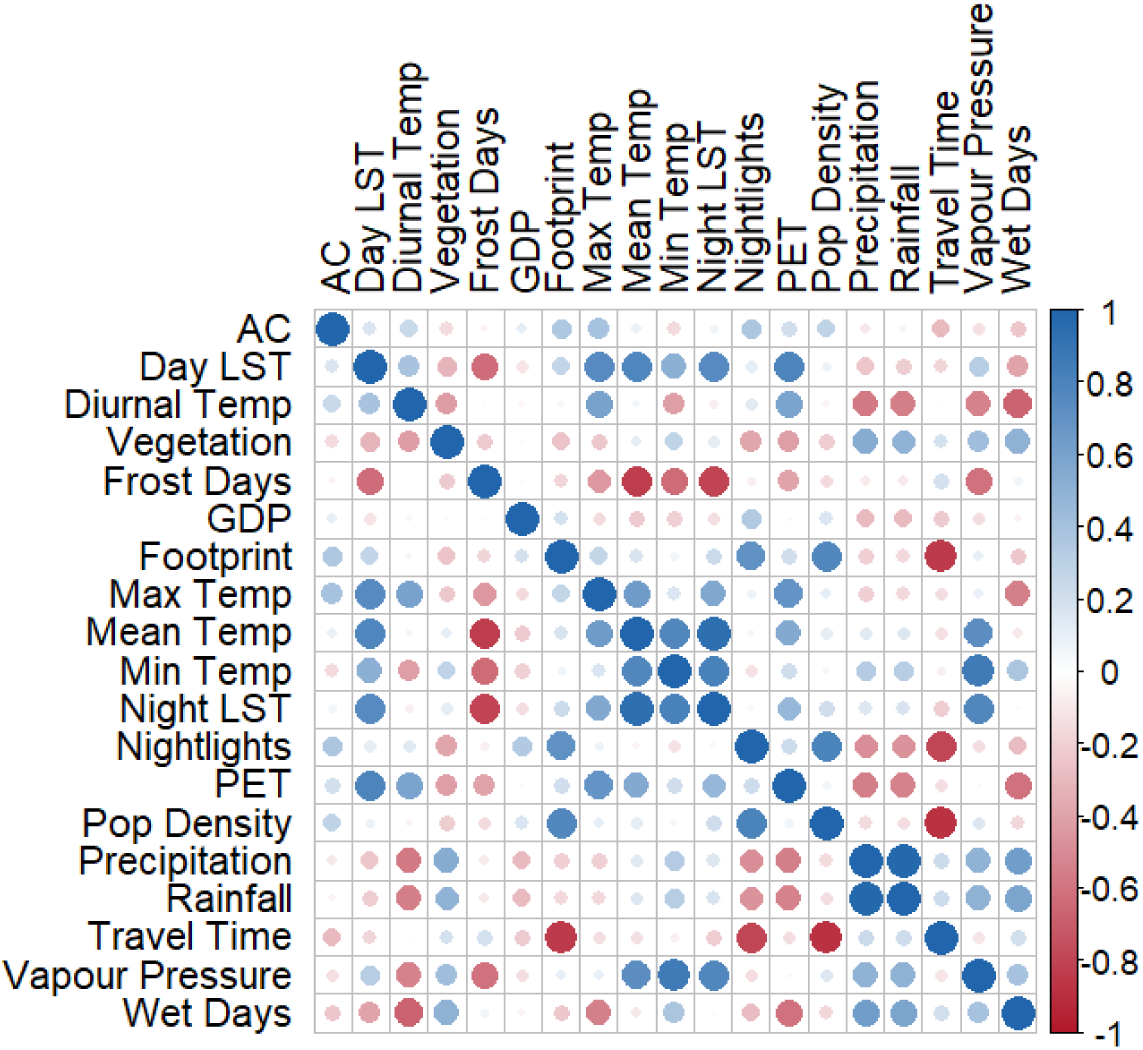
Pairwise correlation matrix of all features in the model.

### Model results

For the elastic net model, the optimal hyperparameters were *α* = 0.1 and *λ* < 0.0001, where alpha controls the balance between ridge and lasso penalisation, and lambda determines the shrinkage applied to regression coefficients. Under 10-fold cross validation, this model achieved a MAE of 0.14. For the decision tree model, the optimal maximum tree depth was 10 and achieved a MAE of 0.12. The random forest model had a MAE of 0.08 with 4 predictors randomly considered at each split (*mtry* = 4) The random forest outperformed the other methods and was therefore selected for further hyperparameter tuning and refinement.

The optimised random forest, after additional hyperparameter tuning and repeated cross-validation, achieved a MAE of 0.08, root mean squared error (RMSE) of 0.13 and an *R*^2^ of 0.73 under repeated 10-fold cross validation (*mtry* = 9, *minimum node size* = 25). Model performance was stable across a wide range of hyperparameter settings, suggesting that predictive accuracy was robust to tuning choices.

To assess model generalisability across countries, we additionally implemented a leave-country-out cross validation. This model achieved a MAE of 0.09, RMSE of 0.13 and *R*^2^ of 0.24, indicating that the model performance varied when predicting in countries not represented in the training data (*mtry* = 3, *minimum node size* = 100). This also reflects how heterogeneous the socioeconomic and environmental conditions are across countries. A summary of results is shown in Table 1. Observed vs predicted plots showed strong alignment along the 45° line, indicating unbiased predictions (Figure 5), though the scatter around this line is considerable.

**Table 1:** Model performance summary.

| | Model | RMSE | $R^2$ | MAE |
| --- | --- | --- | --- | --- |
| Simple models | Elastic net | 0.189 | 0.396 | 0.141 |
|  | Decision tree | 0.177 | 0.471 | 0.118 |
|  | Random forest | 0.127 | 0.728 | 0.078 |
| Hyper Parameter Tuned | Refined random forest | 0.127 | 0.733 | 0.078 |
|  | Leave country out random forest | 0.128 | 0.243 | 0.091 |

Although RMSE was similar across the random forest models, differences in MAE and *R*^2^ reflect how prediction errors vary by clusters. RMSE squares errors before averaging and was dominated by many near zero values, whereas MAE captures the typical prediction accuracy of most clusters. *R*^2^ was strongly influenced by higher concentration of clusters with very low AC prevalence, especially when compared in different countries. Because AC prevalence is highly skewed with many clusters having near zero values, squared errors from these clusters are very small, which is why the RMSE did not vary considerably between models. Contrary to this, MAE is more sensitive to differences in prediction accuracy across clusters, and therefore the magnitude of prediction errors across clusters is reflected by the changes in MAE.

### Spatial predictions of AC ownership in 2023

Using the optimised random forest model, we generated a continent-wide prediction of household AC ownership across Africa and Asia for the year 2023. Predictions were made at a 5km spatial resolution by applying the fitted model to the raster stack of environmental, climatic and socioeconomic covariates compiled for 2023. These predictions constitute the first spatially explicit, high-resolution map of AC prevalence across low- and middle-income countries in Africa and Asia and are mapped in Figure 3. These predictions can be downloaded at https://doi.org/10.6084/m9.figshare.34022040. Additionally, we plotted regional maps of these predictions which can be downloaded at https://doi.org/10.6084/m9.figshare.34031325.

### Summary statistics of model results

During the period from 2014 to 2023, an estimated average of 2.38 billion people lived in areas where annual maximum temperature reached at least 35° *C*. Of these people an average 1.78 billion (± 0.02 billion) were predicted to lack AC, representing 74.7 % of the population. At a higher threshold of 40° *C*, the average exposed population over the ten-year period was 1.01 billion (± 0.03 billion) out of which 0.69 billion did not have access to AC representing 68.6 % of the population. A full table of these yearly results can be seen in Table 2.

**Table 2:** Summary of population counts in billions of total numbers of people exposed to 35 and 40 degrees Celsius, and the number of people and percentage of those that lack access to air conditioning in these extreme temperature regions.

| Year | Population Exposed to 35° C |  | Population Exposed to 40° C |  |
| --- | --- | --- | --- | --- |
|  | Total | No AC (n(%)) | Total | No AC (n(%)) |
| 2014 | 2.37 | 1.77 (74.9%) | 1.03 | 0.69 (67.2%) |
| 2015 | 2.33 | 1.73 (74.5%) | 1.07 | 0.75 (69.5%) |
| 2016 | 2.48 | 1.86 (74.9%) | 1.19 | 0.82 (68.8%) |
| 2017 | 2.44 | 1.83 (74.8%) | 1.10 | 0.75 (68.6%) |
| 2018 | 2.22 | 1.65 (74.1%) | 1.09 | 0.74 (67.8%) |
| 2019 | 2.46 | 1.84 (74.8%) | 1.05 | 0.71 (67.9%) |
| 2020 | 2.31 | 1.72 (74.3%) | 0.96 | 0.66 (68.5%) |
| 2021 | 2.34 | 1.75 (74.7%) | 0.84 | 0.59 (69.3%) |
| 2022 | 2.43 | 1.81 (74.5%) | 1.08 | 0.73 (67.6%) |
| 2023 | 2.45 | 1.84 (74.9%) | 0.68 | 0.48 (71.0%) |

### Extended analysis on leave country out cross validation

The correlation coefficient between observed and predicted AC prevalence between countries at national level was 0.30 showing a weak relationship. However, within-country correlations at cluster level had a mean of 0.48, ranging from 0.16 to 0.76. This suggests that the model had a moderate ability to rank locations within countries despite the poor overall prediction of AC prevalence. Predicted values also had a lower within-country variability (standard deviation = 0.06) than observed values (standard deviation = 0.11) due to the model trying to smooth out spatial variation. As a result, clusters with high AC prevalence tend to be underpredicted, while those with low prevalence are overpredicted. This is a common problem in machine learning spatial prediction. For example, in Tajikistan, the model captured within-country spatial ranking with a correlation of 0.76 between observed and predicted values but had poor calibration of absolute AC prevalence with an observed mean of 0.28 and predicted mean of 0.11. Other countries like Egypt had both poor calibration (observed mean = 0.06 and predicted mean = 0.37) and within-country correlation of 0.16. This shows that the model could not extrapolate country-level mean prevalence to unseen countries. A full table of these correlations can be found in Figure 6 and Table 3.

### Patterns of AC distribution

Predicted AC ownership displayed spatial variation across Africa and Asia, aligning with differences in climate, urbanisation and socioeconomic development. Much of sub-Saharan Africa is predicted to have very low predicted AC prevalence, with most areas estimated to have less than 20% prevalence in a cluster, especially in Central and Western Africa. In contrast, several regions of South Asia have very high predicted prevalence especially northern and central India and Pakistan with values exceeding 40% per cluster in some areas. Moderate levels of predicted AC prevalence were predicted in parts of Southeast Asia around densely populated and developed areas. Some coastal regions of North Africa were predicted to have moderately higher predicted AC prevalence compared to the rest of the continent. These patterns are mirrored by the relationships observed in the scatterplot and correlation analysis where variables like night LST, GDP, population density and nightlights were positively associated with AC prevalence. Urban areas consistently showed higher predicted values, showing the role of electricity infrastructure and economic activity in enabling access to cooling. Areas with high average temperatures also had greater predicted AC prevalence.

**Figure 2:**
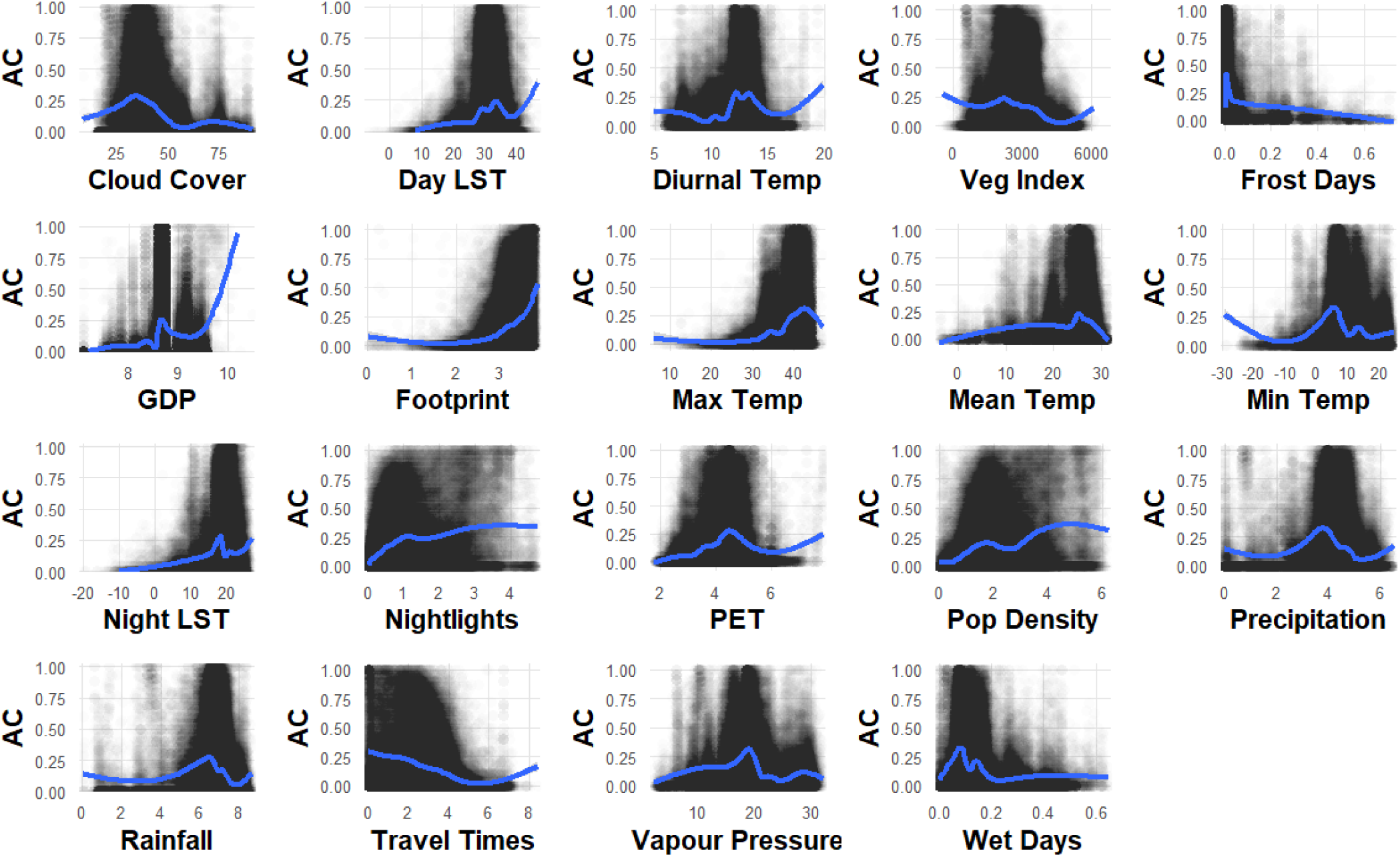
Scatterplots showing the relationships between each feature in the model and air conditioner (AC) prevalence. The features include cloud cover (%), average day land surface temperature (° C), diurnal temperature range (° C), enhanced vegetation index, proportion of frost days in a year, log gross domestic product (GDP), log global human footprint index, maximum temperature (° C), mean temperature (° C), minimum temperature (° C), average nighttime land surface temperature (° C), log nightlights, potential evapotranspiration (mm), log population density (persons per km^2^), log precipitation (mm), log rainfall (mm), log travel times (minutes), vapour pressure (hPa) and proportion of wet days in a year (0-1). Variables labelled log were transformed using the natural logarithm of the original value plus one [ln(x + 1)]. The temporal resolution is yearly.

**Figure 3:**
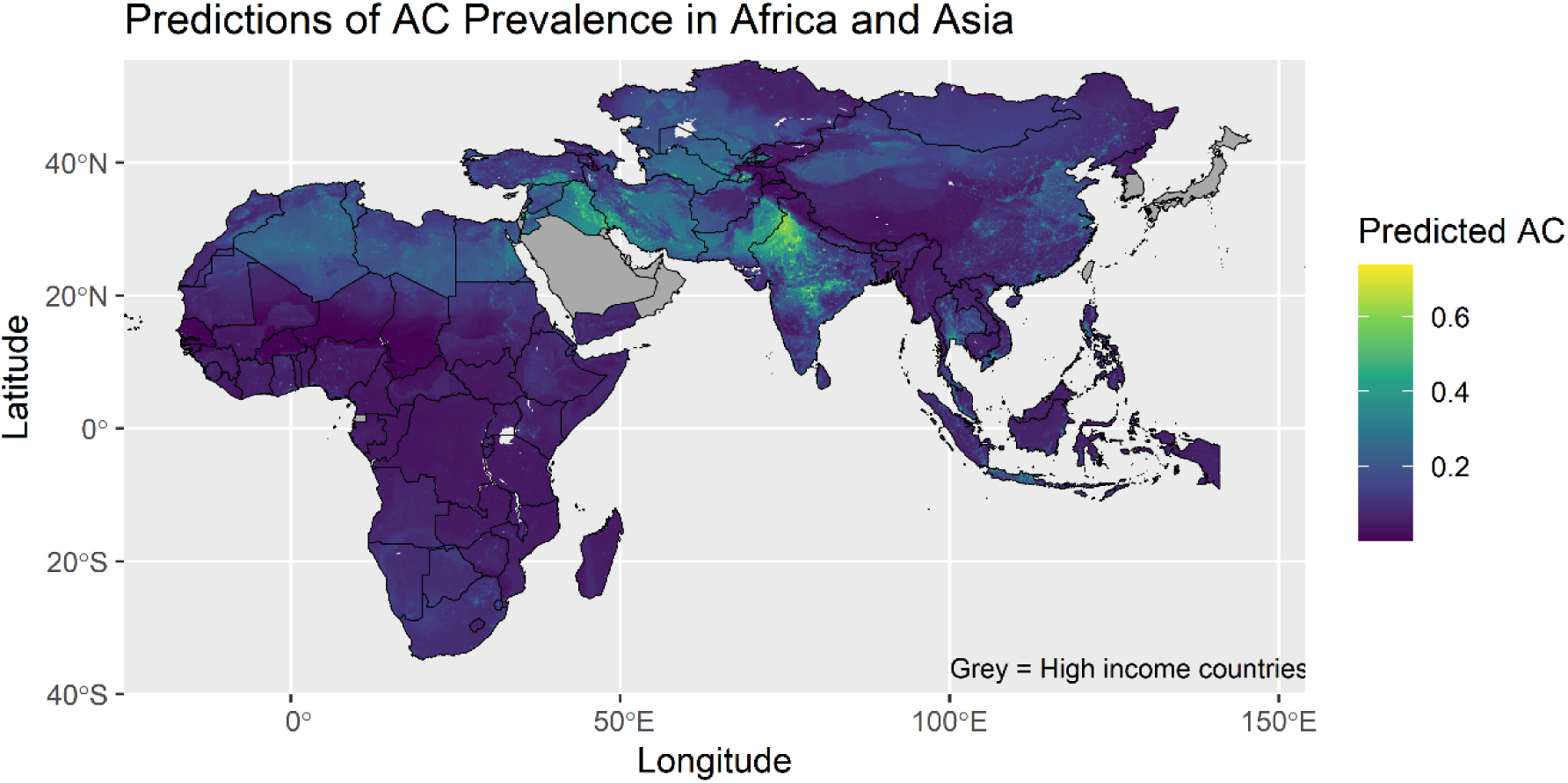
Spatial predictions of Air Conditioner prevalence in Africa and Asia at 5km resolution in the year 2023. Grey countries represent high income countries.

## Discussion

This study makes a novel contribution by producing the first high-resolution (5 km) continental scale map of household AC prevalence across Africa and Asia. Overall, the spatial predictions of AC prevalence in Africa and Asia in the year 2023 are consistent with established disparities in income, infrastructure and heat exposure. It shows that AC remains concentrated in wealthier, hotter and more urbanised regions leaving vulnerable populations in deprived areas and lower income families without access to cooling highlighting the scale of the “adaptation cooling gap” across Africa and Asia.

Our high-resolution predictions enable future assessments of heat-related risks and energy demand as well as providing a baseline to monitor trajectories of AC uptake under various climate and socioeconomic changes. However, predictions for countries absent in the DHS training data should be interpreted with caution given the reduced performance in leave-country-out cross-validation. By combining nationally representative household survey data with geospatial climatic and socioeconomic covariates and applying machine learning models, this study advances knowledge in cooling access in regions where extreme heat is intensifying but data availability is limited.

This work is related to the United Nations Sustainable Development Goals, particularly the 10^th^ goal of reducing inequalities and the 7^th^ and 11^th^ goals of ensuring access to clean energy and making settlements more inclusive and sustainable. Our work contributes to these goals via the AC prevalence map which provides evidence that can inform equitable heat adaptation planning and guide allocation of resources for planning and development.

### Context of prior research

Our approach builds on recent efforts to quantify cooling access and needs. Ahn and Uejio (2022) demonstrated that random forests can predict AC ownership at fine spatial scales, but their analysis was confined to California using property level data. By extending this approach across two continents and integrating household survey data linked to environmental and socioeconomic predictors, this study allows predictions in areas where property-level data is not available. Similarly, Biardeau (et al. 2019) highlighted global disparities by linking population data with Cooling Degree Days (CDDs), showing that countries like India, Nigeria and Indonesia are already experiencing higher cooling demand than the United States. However, they relied on potential cooling demand and used aggregate climate proxies which did not include socioeconomic or infrastructural drivers of AC. Contrary to this, our analysis integrates GDP, population density, night lights and various climatic factors aggregated annually with DHS surveys to model cooling access. This allows spatial predictions that reflect drivers of AC demand.

This study also connects the concept of the adaptation cooling deficit explained by Davis (et al. 2021), which captures the mismatch between populations exposed to heat stress and those with access to cooling. Their analysis focused on relationships between income and climate on a national level in emerging economies whereas our results show how this deficit exhibits itself within countries and highlights urban-rural divisions and regional disparities. Our findings are consistent with projections by (Falchetta et al. 2024) who forecasts that rising AC adoption will magnify the existing global energy inequality. Our analysis provides a baseline for 2023 AC prevalence.

### Spatial patterns

The predicted spatial distribution of AC ownership in this study reflects both socioeconomic and climatic gradients. These factors explain the much higher predicted prevalence in large metropolitan areas including capitals relative to their rural surroundings. Rural areas with limited electricity access were predicted to have low ownership.

Climatic variables also play a major role with higher mean and maximum temperatures, greater diurnal temperature ranges and intense nighttime heat all linked to increased AC prevalence. Conversely, areas with cooler conditions and frequent frost days showed low predicted ownership. These patterns highlight barriers in rural and low-income areas where affordability, infrastructure and electricity reliability stays inadequate, leaving large populations exposed despite harsh climatic conditions. While the aim of our analysis is explicitly predictive, the fact that these relationships match reasonable causal expectations provides a sense check for the models.

### Strengths of the study

A key strength of this study is the use of geolocated DHS household surveys across 24 countries matched with high resolution environmental and socioeconomic rasters aligned to survey years. The modelling framework used random forests, which are known for strong predictive performance and flexibility with complex non-linear relationships involving skewed variables. A rigorous cross-validation scheme was applied using repeated k-fold and leave-country-out designs to test for generalisability and robustness. The resulting 5km resolution map of predicted AC ownership in 2023 provides a ready visual dataset that can be applied by policy makers in adaptation and energy planning. The publicly available raster data of our predictions can be used in further analyses.

### Limitations

Despite these strengths, there are some limitations of this study. Firstly, generalisability across countries was weak as seen under the leave-country-out cross-validation due to the heterogeneous socioeconomic and infrastructural differences in countries. Secondly, DHS coverage itself was sparse with only 24 countries included, which leaves major gaps in regions like East Asia and the Middle East. Additionally, proxies such as GDP and nightlights imperfectly capture household income and electricity access. GDP is also recorded annually at the country level and therefore does not capture household level income. Further, DHS cluster GPS are deliberately displaced to protect confidentiality which introduces spatial noise.

This modelling framework could be extended by applying a spatial-temporal modelling approach to allow trends in AC adoption over time. The model could also benefit from external validation sources such as appliance sales data.

While DHS provides a unique combination of household appliance ownership and geographic coordinates, there are some surveys that exist which either record AC ownership without corresponding GPS coordinates or include spatial information without appliance ownership data. These datasets could potentially be included in future work. The predictions presented in this study are an initial spatial estimate of AC prevalence made in the year 2023, but the dynamic nature of AC adoption means that patterns may change over time. Additionally, given that AC units typically last for decades, models that account for the stock-and-flow nature of these assets might be appropriate (Tan et al. 2026).

The highly skewed nature of the distribution of AC prevalence across clusters was reflected in our model results. It was observed that the RMSE remained relatively similar across models and this is because most clusters in the dataset have very low or zero AC ownership, with very few clusters having substantially higher prevalence. Since RMSE is calculated by squaring the prediction errors, it is driven by very few large errors in this model, and many near zero observations contributing very little to the overall RMSE. In contrast, the MAE measures the average magnitude of prediction errors without squaring them and therefore reflects the typical prediction across clusters making it more sensitive to differences in model performance. Lastly *R*^2^ depends on the proportion of variance explained and is strongly influenced by differences in AC prevalence between countries. When models were evaluated using leave-country-out cross-validation, the differences in between-country information led to a large decrease in explained variance. Additional analysis suggested that while estimation of country-level mean AC prevalence was poor, the model retained a moderate ability to rank locations in unseen countries. This suggests that the model better captures broad spatial gradients and patterns rather than absolute national prevalence. Overall, given the skewed distribution of AC prevalence across clusters, MAE was considered the most informative indicator of predictive performance as it highlights the magnitude of prediction error without being influenced unproportionally by extreme values.

Leave-country-out cross-validation evaluates the ability for the model to transfer learned relationships to entirely unseen countries. The weighted random forest model is trained on all 24 DHS countries and then generates predictions across all LMIC regions in Africa and Asia. This tells us more about the spatial structures and relationships learned by the model from available survey data. Therefore, the model’s aim is to understand how the model behaves when predicting across the continental raster surface, and how reliable these predictions are. Comparing this to the leave-country-out framework helps us distinguish between the model’s ability to reconstruct observed spatial variation in represented countries and its ability to extrapolate to countries where no AC survey data are available.

### Applications and policy relevance

Despite the drawbacks, the outputs of this study have direct applications for adaptation and sustainability policy. The map identifies areas with critical adaptation cooling gaps, showing that AC access is concentrated in wealthier, urban and hotter regions while rural and low-income populations remain largely excluded. These insights can support equitable targeting of subsidies, development of electricity lines to make them more accessible, as well as anticipating future demand of electricity and associated emissions. It also provides resources to epidemiologists enabling further observational studies assessing the protective and adverse health impacts of AC, as well as industrialists who might use this information to target localised allocation of resources to areas in need of AC.

These findings can also be interpreted within the context of global energy poverty. Recent evidence highlighted by Our World in Data (Ritchie 2026) shows that in many low-income countries the availability of residential electricity is not sufficient to power a single room air conditioner for one hour a day. For example, in India the average use of electricity per person would only allow 44 minutes of AC use, 13 minutes in Nigeria and as little as 4 minutes in South Sudan. This illustrates how adaptation inequalities are not only caused by AC affordability but also limited access to electricity. As a result, people living in some of the hottest regions globally are unable to access cooling technologies despite the continuous rising temperatures.

### Future directions

Future work can look at building on this model by incorporating richer socioeconomic indicators at household level, wealth indices, electricity tariffs as well as national data on AC sales for external validation to strengthen predictions. Geographically, extending the analysis by including more countries beyond Africa and Asia would provide a global picture. Finally, integrating predicted AC prevalence with health outcome data such as heat-related mortality would facilitate direct epidemiological applications and strengthen evidence for climate adaptation policies.

## Conclusion

By developing the first continental scale machine learning model, we provide both a methodological advancement and policy-relevant evidence base showing AC prevalence in LMICs across Africa and Asia.

## Data Availability

The resulting dataset of predicted air conditioner prevalence in low- and middle-income countries has been made publicly available on https://doi.org/10.6084/m9.figshare.34022040

https://catalogue.ceda.ac.uk/uuid/715abce1604a42f396f81db83aeb2a4b/

https://doi.org/10.5067/MODIS/MOD11C3.061

https://zenodo.org/records/13943886

https://doi.org/10.6084/M9.FIGSHARE.16571064

https://doi.org/10.1038/s41597-020-0453-3

https://doi.org/10.1080/01431161.2017.1342050

https://doi.org/10.5258/SOTON/WP00845

https://data.malariaatlas.org/maps?layers=Accessibility:201501_Global_Travel_Time_to_Cities,Malaria:202508_Global_Pf_Parasite_Rate

https://dhsprogram.com/data/

## Supplementary material

**Figure 4:**
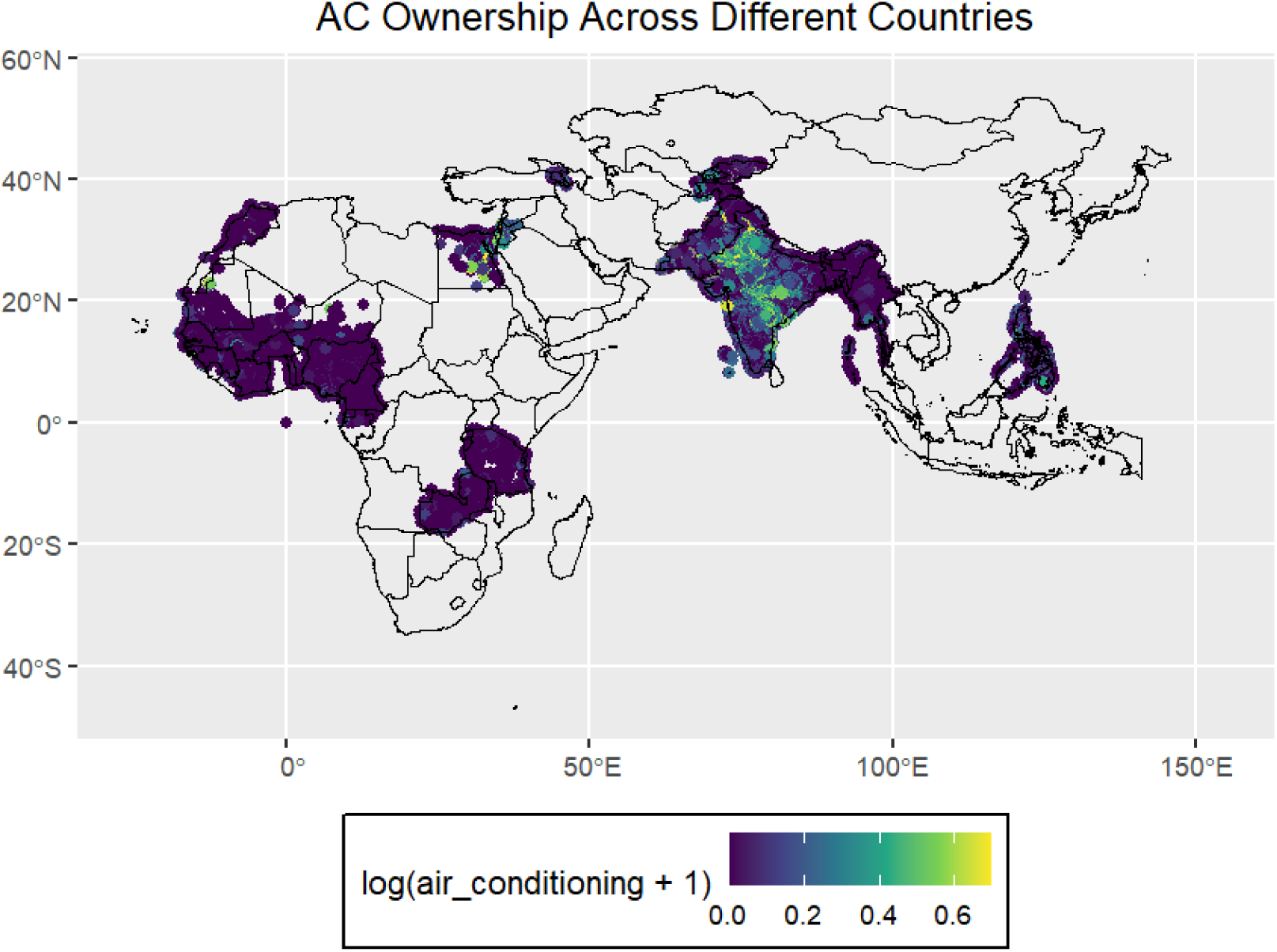
Raw DHS data of Air Conditioner Ownership across 24 countries

**Figure 5:**
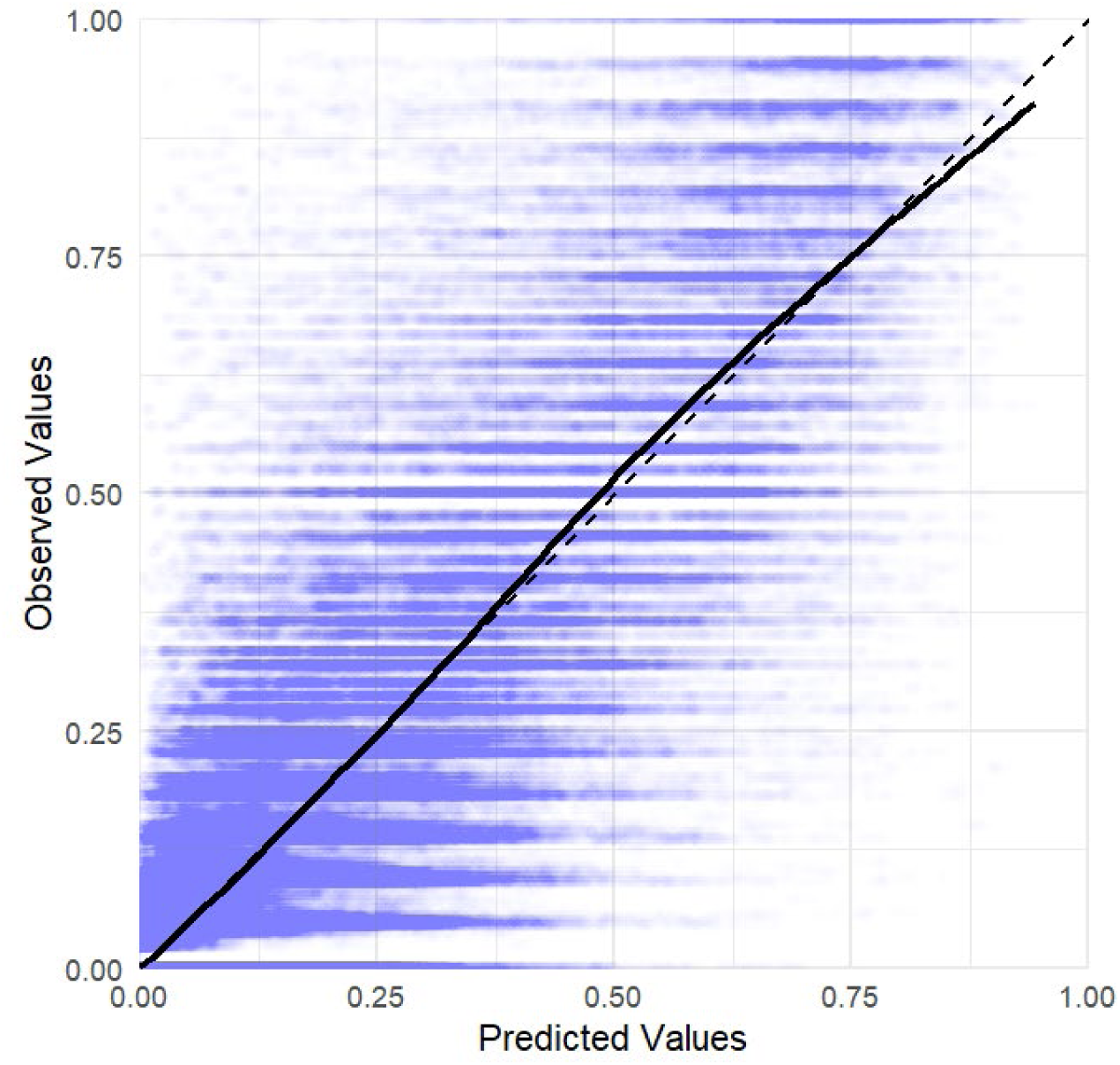
Observed vs Predicted plot of AC prevalence in the refined weighted random forest model.

**Figure 6:**
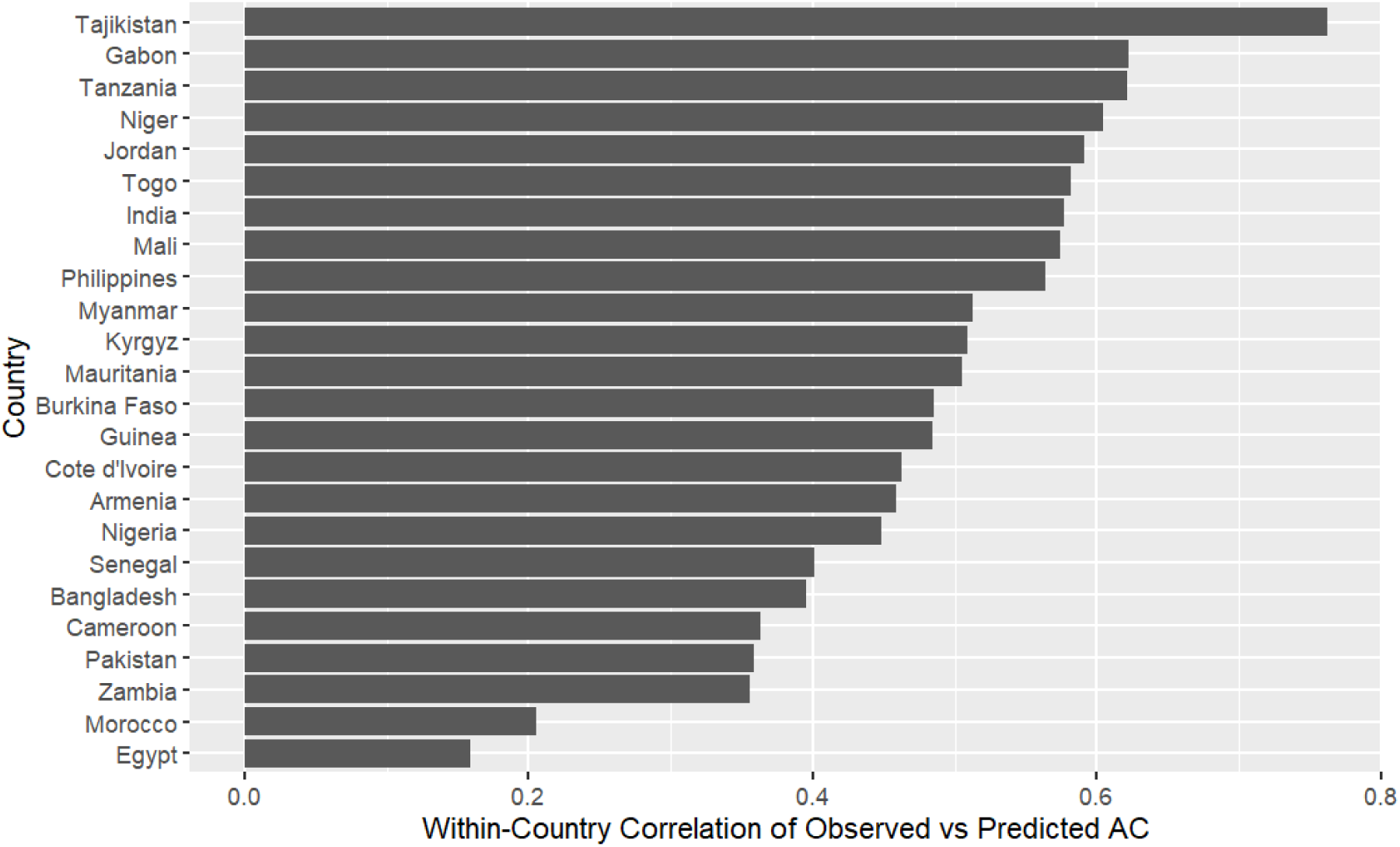
Within country correlation of observed vs predicted AC prevalence at DHS cluster level for the leave country out cross validation.

**Table 3:**
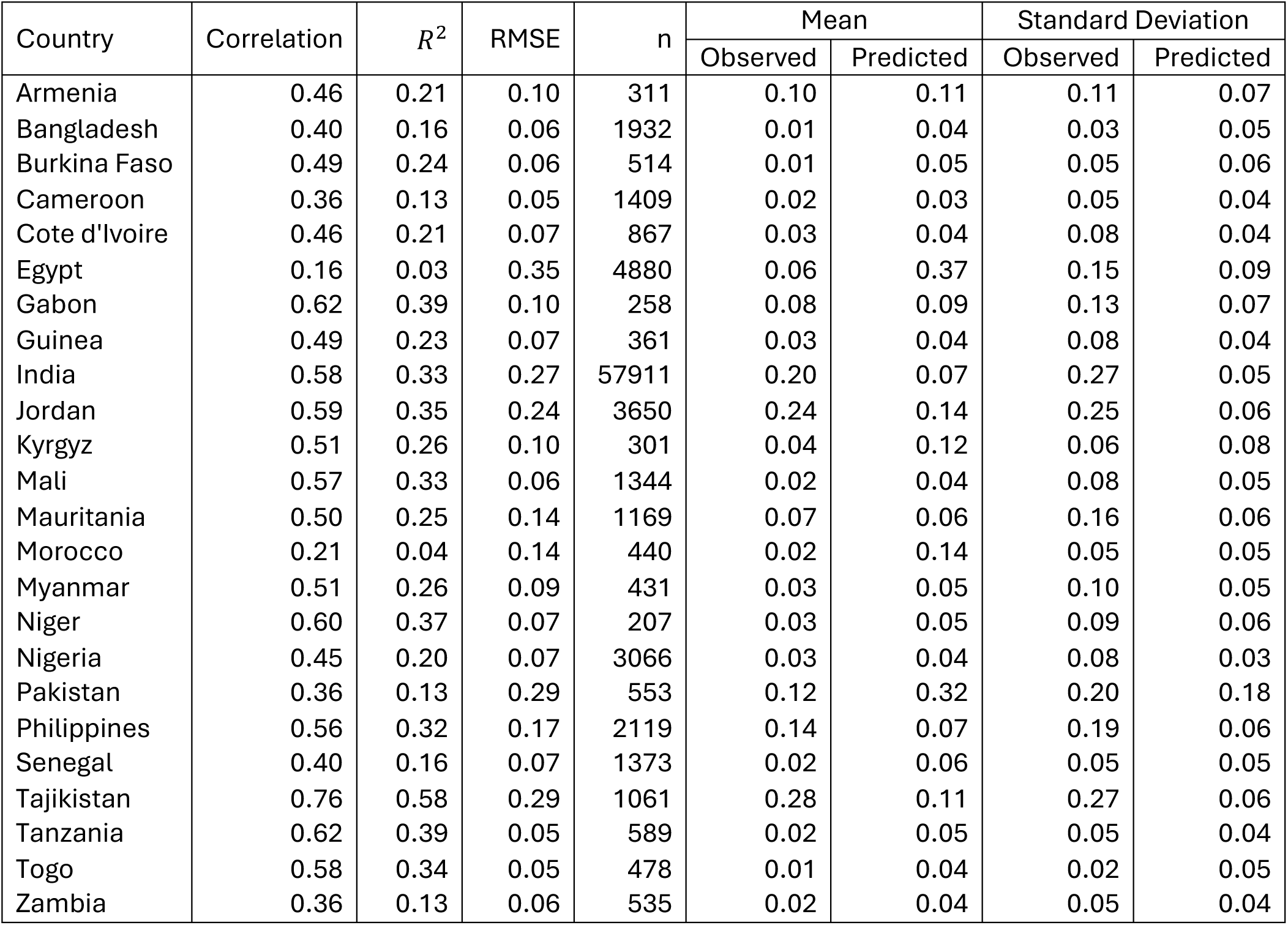
Country specific performance metrics from leave-country-out cross-validation random forest model predicting household AC prevalence. Correlation represents within-country correlation between observed and predicted DHS cluster level AC prevalence. Observed and predicted means and standard deviation summarise the country level distributions of AC prevalence. N denotes the number of DHS clusters within each held out country.

| Country | Correlation | $R^2$ | RMSE | n | Mean | | Standard Deviation | |
| --- | --- | --- | --- | --- | --- | --- | --- | --- |
|  |  |  |  |  | Observed | Predicted | Observed | Predicted |
| Armenia | 0.46 | 0.21 | 0.10 | 311 | 0.10 | 0.11 | 0.11 | 0.07 |
| Bangladesh | 0.40 | 0.16 | 0.06 | 1932 | 0.01 | 0.04 | 0.03 | 0.05 |
| Burkina Faso | 0.49 | 0.24 | 0.06 | 514 | 0.01 | 0.05 | 0.05 | 0.06 |
| Cameroon | 0.36 | 0.13 | 0.05 | 1409 | 0.02 | 0.03 | 0.05 | 0.04 |
| Cote d'Ivoire | 0.46 | 0.21 | 0.07 | 867 | 0.03 | 0.04 | 0.08 | 0.04 |
| Egypt | 0.16 | 0.03 | 0.35 | 4880 | 0.06 | 0.37 | 0.15 | 0.09 |
| Gabon | 0.62 | 0.39 | 0.10 | 258 | 0.08 | 0.09 | 0.13 | 0.07 |
| Guinea | 0.49 | 0.23 | 0.07 | 361 | 0.03 | 0.04 | 0.08 | 0.04 |
| India | 0.58 | 0.33 | 0.27 | 57911 | 0.20 | 0.07 | 0.27 | 0.05 |
| Jordan | 0.59 | 0.35 | 0.24 | 3650 | 0.24 | 0.14 | 0.25 | 0.06 |
| Kyrgyz | 0.51 | 0.26 | 0.10 | 301 | 0.04 | 0.12 | 0.06 | 0.08 |
| Mali | 0.57 | 0.33 | 0.06 | 1344 | 0.02 | 0.04 | 0.08 | 0.05 |
| Mauritania | 0.50 | 0.25 | 0.14 | 1169 | 0.07 | 0.06 | 0.16 | 0.06 |
| Morocco | 0.21 | 0.04 | 0.14 | 440 | 0.02 | 0.14 | 0.05 | 0.05 |
| Myanmar | 0.51 | 0.26 | 0.09 | 431 | 0.03 | 0.05 | 0.10 | 0.05 |
| Niger | 0.60 | 0.37 | 0.07 | 207 | 0.03 | 0.05 | 0.09 | 0.06 |
| Nigeria | 0.45 | 0.20 | 0.07 | 3066 | 0.03 | 0.04 | 0.08 | 0.03 |
| Pakistan | 0.36 | 0.13 | 0.29 | 553 | 0.12 | 0.32 | 0.20 | 0.18 |
| Philippines | 0.56 | 0.32 | 0.17 | 2119 | 0.14 | 0.07 | 0.19 | 0.06 |
| Senegal | 0.40 | 0.16 | 0.07 | 1373 | 0.02 | 0.06 | 0.05 | 0.05 |
| Tajikistan | 0.76 | 0.58 | 0.29 | 1061 | 0.28 | 0.11 | 0.27 | 0.06 |
| Tanzania | 0.62 | 0.39 | 0.05 | 589 | 0.02 | 0.05 | 0.05 | 0.04 |
| Togo | 0.58 | 0.34 | 0.05 | 478 | 0.01 | 0.04 | 0.02 | 0.05 |
| Zambia | 0.36 | 0.13 | 0.06 | 535 | 0.02 | 0.04 | 0.05 | 0.04 |

**Table 4:** Summary of variables extracted in raster format, their source and coverage.

| **Variables** | **Source** | **Coverage** |
| --- | --- | --- |
| Cloud cover, diurnal temperature, frost days, mean temperature, potential evapotranspiration, vapour pressure, wet days | CEDA Archive (University of East Anglia Climatic Research Unit et al. 2024) | 2001–2023 |
| Day land surface temperature | EarthData (Wan et al. 2021a) | 2001–2023 |
| Enhanced vegetation index | EarthData (Didan and Barreto 2016) | 2001–2023 |
| Gross domestic product | Zenodo (Kummu et al. 2024) | 2001–2022 |
| Global human footprint | Figshare (Mu et al. 2025) | 2001–2022 |
| Maximum and minimum temperature, precipitation | WorldClim (Harris et al. 2020) | 2001–2023 |
| Night land surface temperature | EarthData (Wan et al. 2021b) | 2001–2023 |
| Nightlights | Earth Observation Group (Elvidge et al. 2017) | 2012–2021 |
| Population density | WorldPop (Bondarenko et al. 2025) | 2001–2020 |
| Rainfall | CHIRPS (Climate Hazards Center 2025) | 2001–2023 |
| Travel times to cities | Malaria Atlas Project (Malaria Atlas Project 2015) | 2015 |
| Urban–rural classification | Figshare (Cattaneo et al. 2021) | Static |

